# Microbiota-Short Chain Fatty Acid Relationships Underlie Clinical Heterogeneity and Identify Key Microbial Targets in Irritable Bowel Syndrome (IBS)

**DOI:** 10.1101/2024.01.31.24302084

**Authors:** Andrea Shin, Yue Xing, Mohammed Rayyan Waseem, Robert Siwiec, Toyia James-Stevenson, Nicholas Rogers, Matthew Bohm, John Wo, Carolyn Lockett, Anita Gupta, Jhalka Kadariya, Evelyn Toh, Rachel Anderson, Huiping Xu, Xiang Gao

## Abstract

**Background:** Identifying microbial targets in irritable bowel syndrome (IBS) and other disorders of gut-brain interaction (DGBI) is challenging due to the dynamic nature of microbiota-metabolite-host interactions. SCFA are key microbial metabolites that modulate intestinal homeostasis and may influence IBS pathophysiology. We aimed to assess microbial features associated with short chain fatty acids (SCFA) and determine if features varied across IBS subtypes and endophenotypes. Among 96 participants who were screened, 71 completed the study. We conducted in-depth investigations of stool microbial metagenomes, stool SCFA, and measurable IBS traits (stool bile acids, colonic transit, stool form) in 41 patients with IBS (IBS with constipation [IBS-C] IBS with diarrhea [IBS-D]) and 17 healthy controls. We used partial canonical correspondence analyses (pCCA), conditioned on transit, to quantify microbe-SCFA associations across clinical groups. To explore relationships between microbially-derived SCFA and IBS traits, we compared gut microbiome-encoded potential for substrate utilization across groups and within a subset of participants selected by their stool characteristics as well as stool microbiomes of patients with and without clinical bile acid malabsorption.

**Results:** Overall stool microbiome composition and individual taxa abundances differed between clinical groups. Microbes-SCFA associations differed across groups and revealed key taxa including *Dorea* sp. CAG:317 and *Bifidobacterium pseudocatenulatum* in IBS-D and *Akkermansia muciniphila* and *Prevotella copri* in IBS-C that that may drive subtype-specific microbially-mediated mechanisms. Strongest microbe-SCFA associations were observed in IBS-D and several SCFA-producing species surprisingly demonstrated inverse correlations with SCFA. Fewer bacterial taxa were associated with acetate to butyrate ratios in IBS compared to health. In participants selected by stool form, we demonstrated differential abundances of microbial genes/pathways for SCFA metabolism and degradation of carbohydrates and mucin across groups. SCFA-producing taxa were reduced in IBS-D patients with BAM.

**Conclusion:** Keystone taxa responsible for SCFA production differ according to IBS subtype and traits and the IBS microbiome is characterized by reduced functional redundancy. Differences in microbial substrate preferences are also linked to bowel functions. Focusing on taxa that drive SCFA profiles and stool form may be a rational strategy for identifying relevant microbial targets in IBS and other DGBI.

## INTRODUCTION

Irritable bowel syndrome (IBS) is a burdensome disorder of gut-brain interaction with an estimated global prevalence rate of 5-10%.^1^ Pathophysiological mechanisms of IBS include disturbances in motility or transit, altered intestinal secretion, impaired intestinal permeability, immune cell reactivity, visceral hypersensitivity, and dysregulated neural signaling and/or central processing.^2^ Despite advancements in the understanding of IBS pathogenesis, diagnostic and therapeutic IBS biomarkers are limited.

Accumulating evidence suggests that the gastrointestinal (GI) microbiome is associated with risk of IBS and may also mediate many of the mechanisms that underlie symptoms including altered motility,^3, 4^ barrier dysfunction,^5^ immune activation,^6, 7^ signaling along the brain-gut axis,^8^ and visceral sensation.^9^ Characterization of microbial composition in IBS has suggested decreased microbial diversity, reduced temporal stability, or changes in the relative abundance of specific bacteria in patients with IBS.^10, 11^ However, these findings have not been sufficiently consistent across studies to establish a clear microbial profile in IBS and gaps in our understanding of the functional microbiome persist. Integrating complementary strategies in the investigation of microbial metabolites will be crucial for gathering actionable insights into the impact of the microbiome in IBS.

Atypical profiles of microbial metabolites including luminal bile acids^12, 13^ and short chain fatty acids (SCFA) have been described in some patients with IBS.^14,15^ Bile acid malabsorption (BAM) is recognized as a mechanistic IBS subtype that can be assessed through several diagnostic methods including measurement of total or primary stool bile acids.^13^ Studies have demonstrated BAM to be associated with physiological traits, symptoms, and quality of life.^16–19^ Recently, researchers have examined microbial contributions to BAM in IBS to report alterations such as enrichment of *Clostridia* bacteria including *C. scindens*,^20^ lower microbial alpha diversity, higher Firmicutes to Bacteroidetes ratio,^19^ and presence of endoscopically visible biofilms correlating with overgrowth of *Escherichia coli* and *Ruminococcus gnavus*.^21^

SCFA are produced by anaerobic fermentation of dietary fibers and resistant starch that enter the colon and regulate intestinal homeostasis and physiology.^22^ Compared to bile acids, the role of SCFA in IBS is less well understood. Studies of stool SCFA in IBS have yielded variable results, which may be related to the heterogeneity of the IBS patient populations and multiple pathways through which SCFA may modulate intestinal physiology. Therefore, while stool SCFA are unlikely to serve as categorical IBS biomarkers, they may provide critical insights into the pathophysiological mechanisms that underlie IBS symptoms or serve as a tool for identifying metabolically relevant microbial targets. Studies^15, 23^ that have assessed stool SCFA in distinct IBS subgroups have reported more consistent associations of stool SCFA with IBS subtypes as well as correlations of stool SCFA with measurable IBS traits such as colonic transit, bowel functions, and bile acid excretion.^23–25^ Despite these reports, the intercorrelation between SCFA and transit time complicates the assessment of whether SCFA profiles represent the metabolic capacity of the resident microbiome. It remains unclear if studying the relationships between the microbiome and excreted SCFA is clinically informative. Recent work^26^ has suggested that individual or keystone taxa, rather than complex ecological communities, could drive changes in SCFA output in response to dietary fiber. Therefore, strategies that isolate major microbial features (or keystone taxa) that shape luminal SCFA may serve as a rational method for selecting functionally relevant microbial targets in patients with IBS. To address these questions, we conducted an in-depth investigation of GI microbiome composition and function, stool SCFA, and IBS endophenotypes defined according to quantitative traits (transit, bile acids, bowel functions) in adults with and without IBS.

## MATERIALS AND METHODS

### Participant Recruitment and Study Design

The study was approved by the Indiana University Institutional Review Board and the protocol registered within ClinicalTrials.gov (NCT02981888). The study was designed as an observational investigation of stool SCFA, stool bile acids, colonic transit, and stool microbiota in adults with and without IBS. We enrolled adults ages 18-65 years of age through the Indiana University Gastroenterology Clinics, the Indiana Clinical and Translational Research Institute Research Registry, and from the local community. We included individuals with IBS with diarrhea (IBS-D) or IBS with constipation (IBS-C) according to Rome IV criteria^27^ and healthy controls with no prior history of GI diseases or symptoms. Detailed eligibility criteria are available in the Supplemental Methods.

### Data Collection

Study eligibility, medications, medical history, and baseline diet using a food frequency questionnaire^28^ were assessed during a screening visit with a study physician. Over a two-week period, data were collected on bowel functions using a standardized bowel pattern diary including the Bristol stool form scale.^29^ All participants submitted a 48-hour stool collection collected during the last 2 days of a 4-day 100 g fat diet, consistent with clinically validated methods for identifying BAM. Specimens were refrigerated during the collection period, returned to the research team on ice, and stored at -80^ο^C.

### Colonic Transit by Radiopaque Markers

Participants underwent assessment of colonic transit time with a previously validated and optimized method using radiopaque markers.^30^

### Stool SCFA and Bile Acids

Frozen aliquots of stool were shipped to the Metabolite Profiling Facility at Purdue University to measure total and individual SCFA concentrations per mg of dry weight by liquid chromatography-mass spectrometry (LC-MS) using published methods^31^ and to the Mayo Clinic Department of Laboratory Medicine and Pathology to measure total and primary stool bile acid levels by high-performance LC-MS through a commercially available, CLIA-approved assay.^16, 32, 33^ Individual SCFA of interest included acetate, butyrate, and propionate as these represent the predominant SCFA produced in humans.^34^

### DNA Extraction, Purification, and Sequencing

Genomic DNA was isolated from stool using the QIAmp® PowerFecal® DNA kit (QIAGEN Inc., Germantown, MD, USA). DNA quality and concentration were on a Qubit fluorometer. Purified DNA underwent library preparation (Nextera XT, Illumina) and paired-end (2x150 bp) sequencing using the NovaSeq v1.5 SP (Illumina, San Diego, CA, USA) to target a sequencing depth of 40M sequences per sample.

### Metagenomic Data Analysis

Metagenomic sequencing reads were quality filtered and processed for taxonomic profiling using MetaPhlAn3. Additive log-ratio (ALR) transformation was applied to analyze differential taxonomic abundances. Based on the identified microbial taxa from each sample, β-diversity indices (e.g., Bray-Curtis dissimilarity) were calculated using the R packages phyloseq and vegan.^35, 36^ Functional profiling was conducted using the HUMAnN3 pipeline annotated by KEGG (Kyoto Encyclopedia of Genes and Genomes) orthogroups (KO).

### Statistical Considerations

We summarized major endpoints of interest: (1) stool microbiome composition, (2) stool SCFA concentrations (total, acetate, propionate, butyrate), total and percent primary stool bile acids, and (3) colonic transit time (overall and segmental). Our primary objective was to quantify the association between microbiome composition and SCFA (microbe-SCFA associations) across clinical groups after controlling for transit and to account for the mechanistic heterogeneity that underlies clinical IBS populations.

We compared microbial taxa abundances between groups and analyzed associations of taxa abundances with IBS endophenotypes/traits (stool SCFA, stool bile acids, and transit) for the collective cohort and within groups. Associations of microbiome composition with group were assessed using general linear regression models (GLM) adjusted for covariates including age, sex, and BMI. Only high abundant species (relative abundance ≥0.1%) that were prevalent in ≥2 specimens within ≥1 clinical group were considered. Partial Canonical Correspondence Analysis (pCCA) conditioned on transit time was employed to quantify the relationship between the microbiome and stool SCFA in all participants and within clinical groups, using the vegan package in R. The significance of the model was assessed through a permutation test with 999 iterations. To identify bacteria whose abundances were strongly associated with the stool SCFA concentrations, we focused on those bacteria scattered along the direction of specific SCFA axis in the biplot, within a 60-degree angle centered around the SCFA axis in both positive and negative directions. The strength of association between the bacteria and the SCFA was ranked by projecting of the bacteria species scores onto each SCFA axis in the biplot. Bacteria with projection scores less than 0.5 were excluded from the subsequent analysis.

For exploratory analyses, we analyzed multivariable associations of microbiome composition with clinical BAM among individuals with IBS-D. Relative abundances were used to examine the associations of gene family/pathway abundances across clinical groups. A manually curated gut-metabolic analysis framework^37^ was applied to examine KO identifiers associated with carbohydrate degradation, SCFA production or metabolism, and mucin degradation using GLM adjusting for covariates. As stool form^38^ has been identified as a strong source of human microbiota variation and closely linked to transit,^39, 40^ we further explored associations of functional potential across groups in a subset of participants who were selected based on stool form features (i.e. those individuals best representing the stool types within their respective clinical group according to bowel diary data) using the Kruskal-Wallis test. For example, we chose the IBS-D and IBS-C participants with the loosest and firmest stool types, respectively, and controls with consistently normal stool types. For all endpoints, missing values were excluded from the analysis for that endpoint.

## RESULTS

### Participant Characteristics

Among 96 volunteers who underwent screening evaluation, 71 completed the study, and 58 participants (Figure 1) with a mean [±SD] age = 35.5 (±13.8) years and mean [±SD] BMI = 26.2 [±7.5] kg/m^2^) were included in the final analysis after excluding those who ineligible (n=16), lost to follow-up (n=9), or and/or did not provide SCFA data (n=13). Baseline clinical characteristics (Table 1) were not significantly different across groups (all p=ns). Comparisons of quantitative traits demonstrated differences in total stool bile acids and transit between IBS-D and control participants and in total stool SCFA and stool acetate) between IBS and controls Table 2, Figures 2-3.

**Figure 1:**
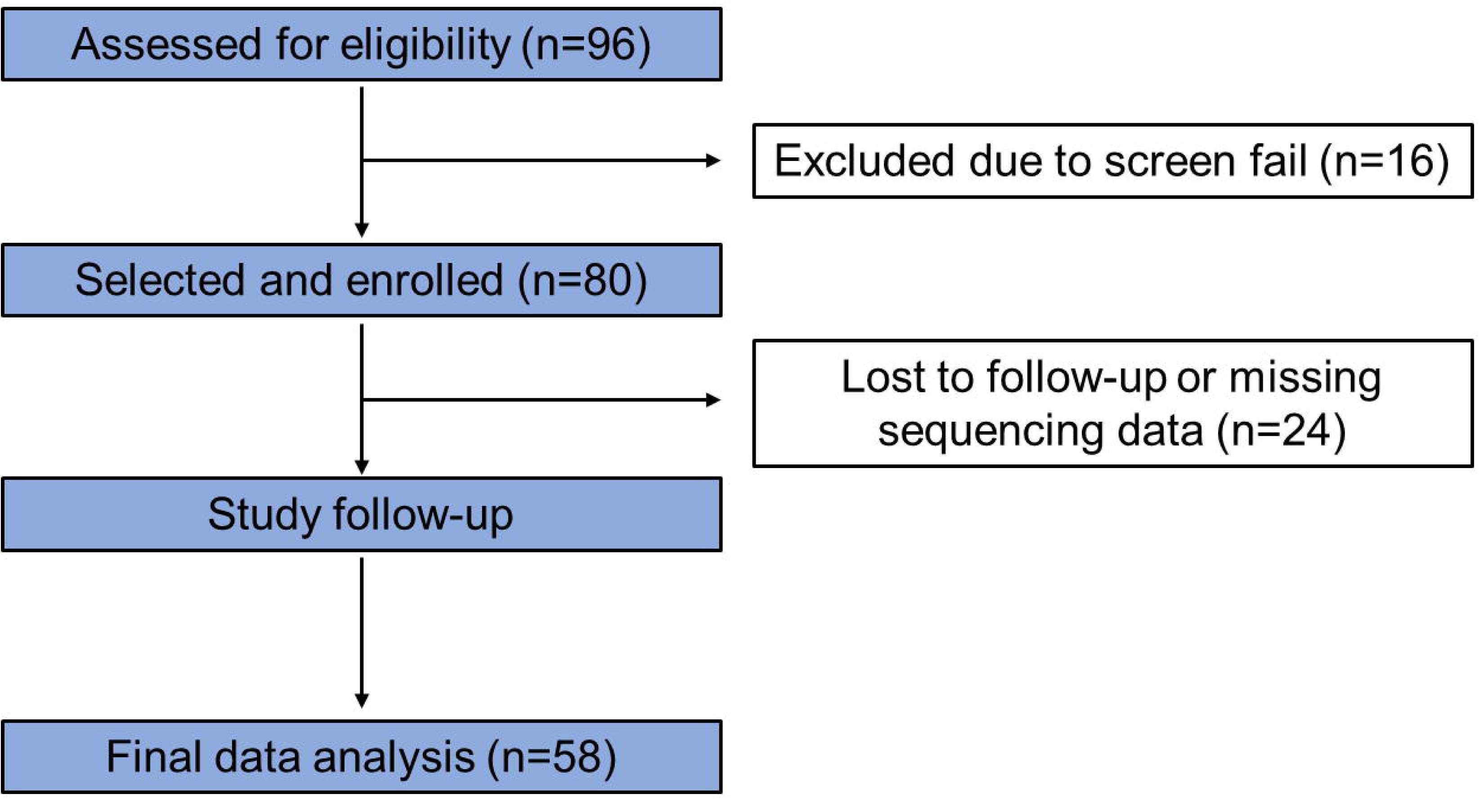
Consort Flow Diagram

**Figure 2:**
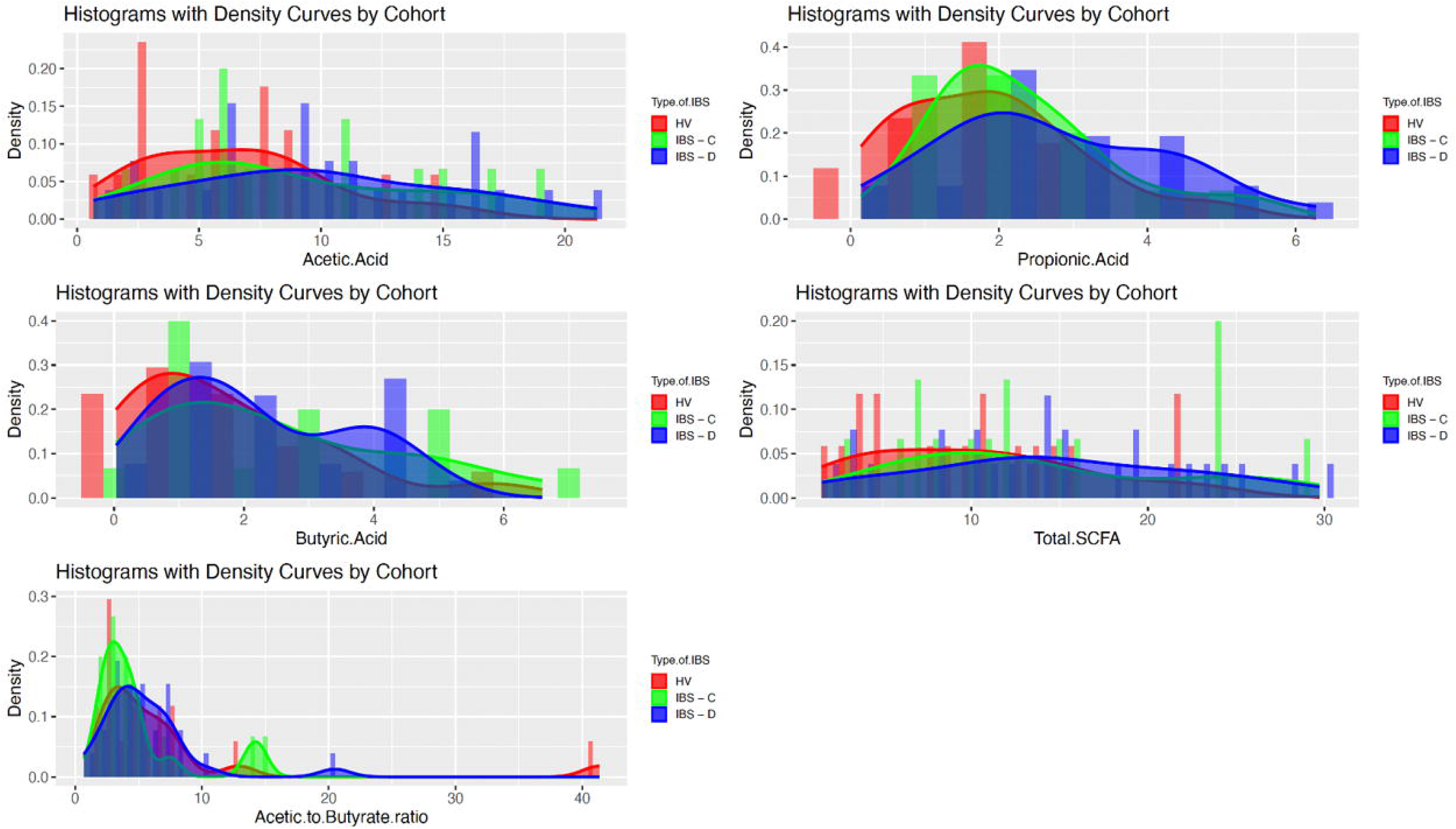
Stool Short Chain Fatty Acid Concentrations in Healthy Volunteers and Participants with Irritable Bowel Syndrome (IBS). *Histograms are shown for total short chain fatty acids, acetate, butyrate, propionate, and acetate to butyrate ratios within clinical groups including healthy volunteers (HV), IBS with constipation (IBS-C), and IBS with diarrhea (IBS-D). Groups are denoted by color (HV = red, IBS-C = green, IBS-D = blue)*.

**Figure 3:**
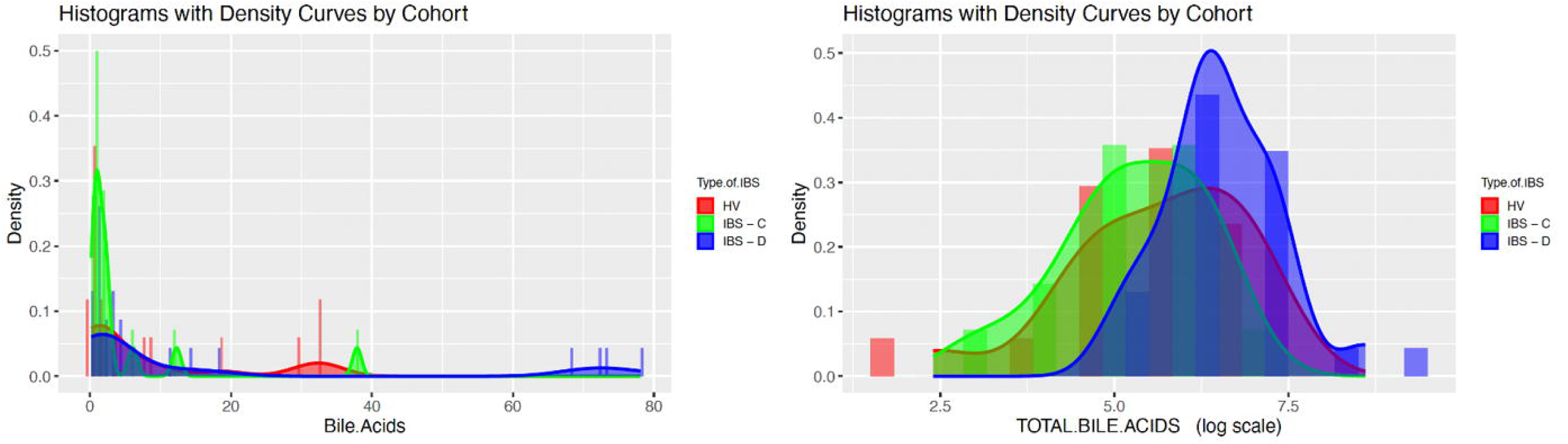
Stool Bile Acids in Healthy Volunteers and Participants with Irritable Bowel Syndrome (IBS). *Histograms are shown for % primary bile acids (bile.acids) and total bile acids within clinical groups including healthy volunteers (HV), IBS with constipation (IBS-C), and IBS with diarrhea (IBS-D). Groups are denoted by color (HV = red, IBS-C = green, IBS-D = blue)*.

**Table 1:** Clinical Characteristics of Patients with Irritable Bowel Syndrome (IBS) and Controls.

| *Data show mean (standard deviation [SD]) except where specified* | **Controls (n=17)** | **IBS-C (n=15)** | **IBS-D (n=26)** |
| --- | --- | --- | --- |
| **Age (years)** | 31.7 (13.6) | 34.3 (10.5) | 38.7 (15.3) |
| **Women, n (%)** | 12 (71%) | 13 (87%) | 18 (70%) |
| **BMI, kg/m<sup>2</sup>** | 26.3 (5.9) | 25.3 (5.2) | 26.7 (9.4) |
| **White, n (%)** | 9 (53%) | 10 (67%) | 23 (88%) |
| **Baseline dietary intake\*** |  |  |  |
| Energy (kcal) | 1946.8 (1025.7) | 2028.4 (1717.3) | 1514.1 (557.4) |
| Total carbohydrates (g) | 219.4 (155.6) | 191.1 (153.7) | 160.1 (62.9) |
| Starch (g) | 104.7 (54.9) | 107.9 (110.9) | 87.7 (46.9) |
| Protein (g) | 100.9 (47.1) | 96.8 (75.3) | 75.1 (23.4) |
| Fat (g) | 77.6 (34.7) | 99.6 (101.9) | 63.8 (30.3) |
| Fiber (g) | 17.9 (20.9) | 14.3 (10.0) | 11.6 (4.1) |

Group comparisons were conducted using the ANOVA F- and Fisher exact tests. IBS-C, IBS with constipation; IBS-D, IBS with diarrhea; \*Diet data missing for two participants with IBS-C and four participants with IBS-D

**Table 2:**
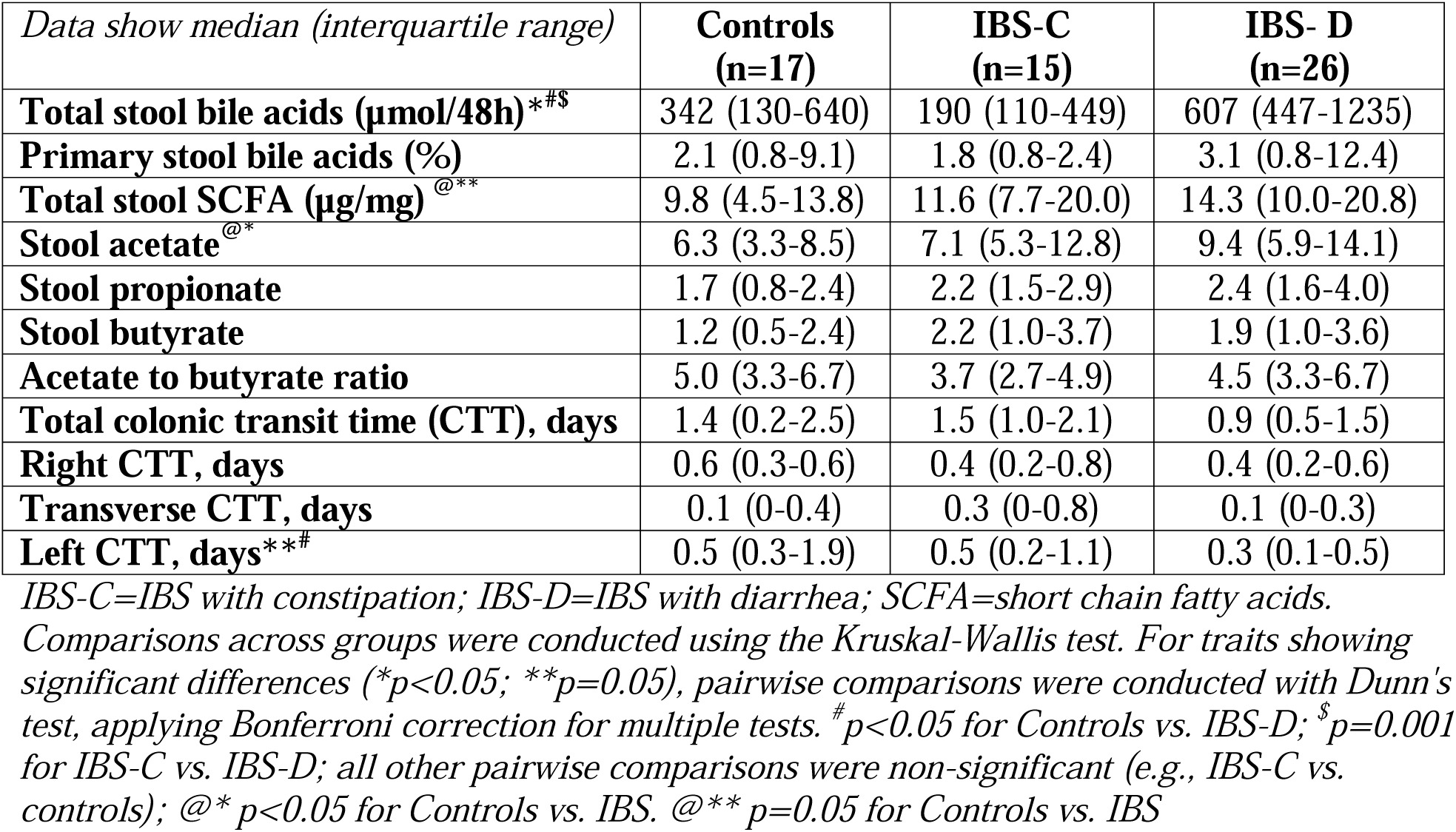
Quantitative Traits in Patients with Irritable Bowel Syndrome (IBS) and Controls.

| *Data show median (interquartile range)* | **Controls**<br>**(n=17)** | **IBS-C**<br>**(n=15)** | **IBS- D**<br>**(n=26)** |
| --- | --- | --- | --- |
| **Total stool bile acids (μmol/48h)\*#** | 342 (130-640) | 190 (110-449) | 607 (447-1235) |
| **Primary stool bile acids (%)** | 2.1 (0.8-9.1) | 1.8 (0.8-2.4) | 3.1 (0.8-12.4) |
| **Total stool SCFA (μg/mg) @\*\*** | 9.8 (4.5-13.8) | 11.6 (7.7-20.0) | 14.3 (10.0-20.8) |
| **Stool acetate @\*** | 6.3 (3.3-8.5) | 7.1 (5.3-12.8) | 9.4 (5.9-14.1) |
| **Stool propionate** | 1.7 (0.8-2.4) | 2.2 (1.5-2.9) | 2.4 (1.6-4.0) |
| **Stool butyrate** | 1.2 (0.5-2.4) | 2.2 (1.0-3.7) | 1.9 (1.0-3.6) |
| **Acetate to butyrate ratio** | 5.0 (3.3-6.7) | 3.7 (2.7-4.9) | 4.5 (3.3-6.7) |
| **Total colonic transit time (CTT), days** | 1.4 (0.2-2.5) | 1.5 (1.0-2.1) | 0.9 (0.5-1.5) |
| **Right CTT, days** | 0.6 (0.3-0.6) | 0.4 (0.2-0.8) | 0.4 (0.2-0.6) |
| **Transverse CTT, days** | 0.1 (0-0.4) | 0.3 (0-0.8) | 0.1 (0-0.3) |
| **Left CTT, days\*\*#** | 0.5 (0.3-1.9) | 0.5 (0.2-1.1) | 0.3 (0.1-0.5) |

IBS-C=IBS with constipation; IBS-D=IBS with diarrhea; SCFA=short chain fatty acids.
Comparisons across groups were conducted using the Kruskal-Wallis test. For traits showing significant differences (\* $p < 0.05$ ; \*\* $p = 0.05$ ), pairwise comparisons were conducted with Dunn's test, applying Bonferroni correction for multiple tests. # $p < 0.05$ for Controls vs. IBS-D; \$ $p = 0.001$ for IBS-C vs. IBS-D; all other pairwise comparisons were non-significant (e.g., IBS-C vs. controls); @\* $p < 0.05$ for Controls vs. IBS. @\*\* $p = 0.05$ for Controls vs. IBS

### Stool microbiome composition differs between IBS and health and between IBS subtypes

Shotgun metagenomic sequencing of stool samples was undertaken to obtain total of 3.1 Gb of sequence data after removal of contaminants with an average of 37.6 million paired-end reads per sample (deposited into NCBI with accession number SUB13882354). Taxonomic classification identified 461 taxa at species level classification. Comparisons of microbial metagenomes based on Bray-Curtis Dissimilarity revealed significant divergence (Supplemental Figure 1) of community distance between groups (p=0.003, R^2^ =0.06) after adjusting for age, BMI, and diet. We compared abundances of individual taxa across groups using the Kruskal-Wallis test to identify 18 unique and differentially abundant taxa. Highest mean relative abundances of 14 taxa (Supplemental Table 1) including *Dorea* sp. CAG:317, *Blautia* sp. CAG:257*, Ruminococcus gnavus,* and *Proteobacteria* bacterium CAG:139 were observed in IBS-D, which have previously been linked to mechanisms such as BAM^20, 21^ and serotonin biosynthesis in patients with IBS-D.^41^ *Lawsonibacter asaccharolyticus* abundance was highest in IBS-C and *Firmicutes* bacterium CAG:83 abundance was highest in controls.

We followed unadjusted analyses with pairwise comparisons of ALR-transformed taxa abundances using covariate-adjusted GLM, focusing on high abundant species, to demonstrate significant differences in 17 pairwise comparisons including 12 unique species (Figure 4) of which 11 exhibited differential abundances of ≥3-fold. Among these taxa, we observed significantly higher abundances of *Dorea* sp. CAG:317 and *Bifidobacterium pseudocatenulatum* in IBS-D compared to IBS-C or controls and higher abundances of *Blautia* sp. CAG:257, and *Proteobacteria* bacterium CAG:139 in IBS-D compared to controls. We found significantly higher abundances of *Clostridium* sp. CAG:58 and lower abundances of *Firmicutes* bacterium CAG:83 in both IBS-D and IBS-C relative to controls, respectively. *Akkermansia muciniphila* and *Prevotella copri* were increased in IBS-C compared to controls. Differences of ≥3-fold in relative abundances among high abundant bacteria were also observed in 151 pairwise comparisons (Supplemental Table 2) but were not statistically significant after adjustment for covariates.

**Figure 4:**
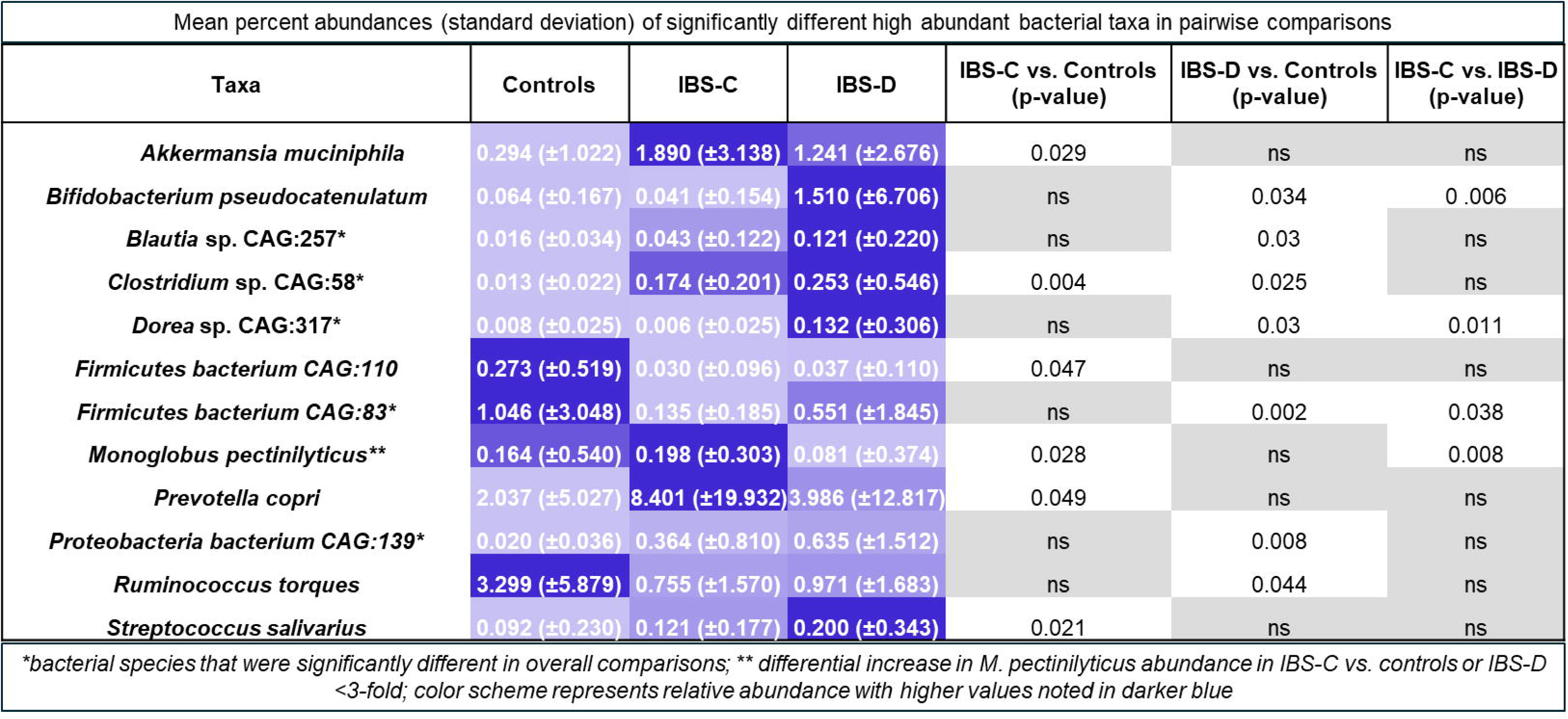
Bacterial Taxa with Differential Abundance of ≥3-fold Between Groups. *Data show mean percent abundances (standard deviation) of significantly different bacterial taxa with* ≥*3-fold differences in pairwise comparisons of clinical cohorts*.

### Microbe-SCFA associations differ between IBS subtypes and controls

We conducted pCCA on microbiome and SCFA data, conditioned on transit time, to quantify relationships between taxa abundances and stool SCFA in the overall cohort and within clinical groups. In the overall cohort, we observed a significant correlation between microbiome abundance matrix and SCFA concentration matrix (p=0.033, *R*^2^ = 0.095). Relationships between microbiome abundances and SCFA concentrations within clinical groups (Supplemental Figure 2) were more pronounced within IBS-D (p=0.015) and less pronounced in HV and IBS-C (p=ns). In the overall cohort, strongest positive associations with total SCFA were observed with *Bacteroides plebeius, Prevotella sp.* CAG:1031, and *Bifidobacterium pseudocatenulatum*.

### Distinct taxa correlate with individual SCFA across clinical subgroups

Keeping in mind that SCFA effects may depend on SCFA type, we ranked bacterial taxa according to their projection scores for individual SCFAs to assess the pattern of associations between bacterial species and SCFA profiles. Bacteria with higher projection scores for individual SCFAs were considered to have stronger associations with those specific SCFAs. Bacteria with projection scores less than 0.5 were excluded from the subsequent analysis. In general, the strength and direction of correlations of individual taxa with acetate, butyrate, and propionate differed according to clinical group (Figure 5). No microbes demonstrated consistent patterns of strong correlations with individual SCFA across all three groups. The greatest number of bacteria with strong associations with SCFA were observed in IBS-D, as evidenced by the larger number of bacteria remaining in the ranked list after filtering. Most microbe-SCFA patterns differed between IBS-D and IBS-C and only a few bacterial species including *Ruminococcus torques*, *Coprococcus comes*, *Clostridium* sp. CAG:299, *Bacteroides eggerthii*, and *Adlercreutzia equolifaciens* demonstrated consistent associations across both IBS subtypes with acetate and butyrate.

**Figure 5:**
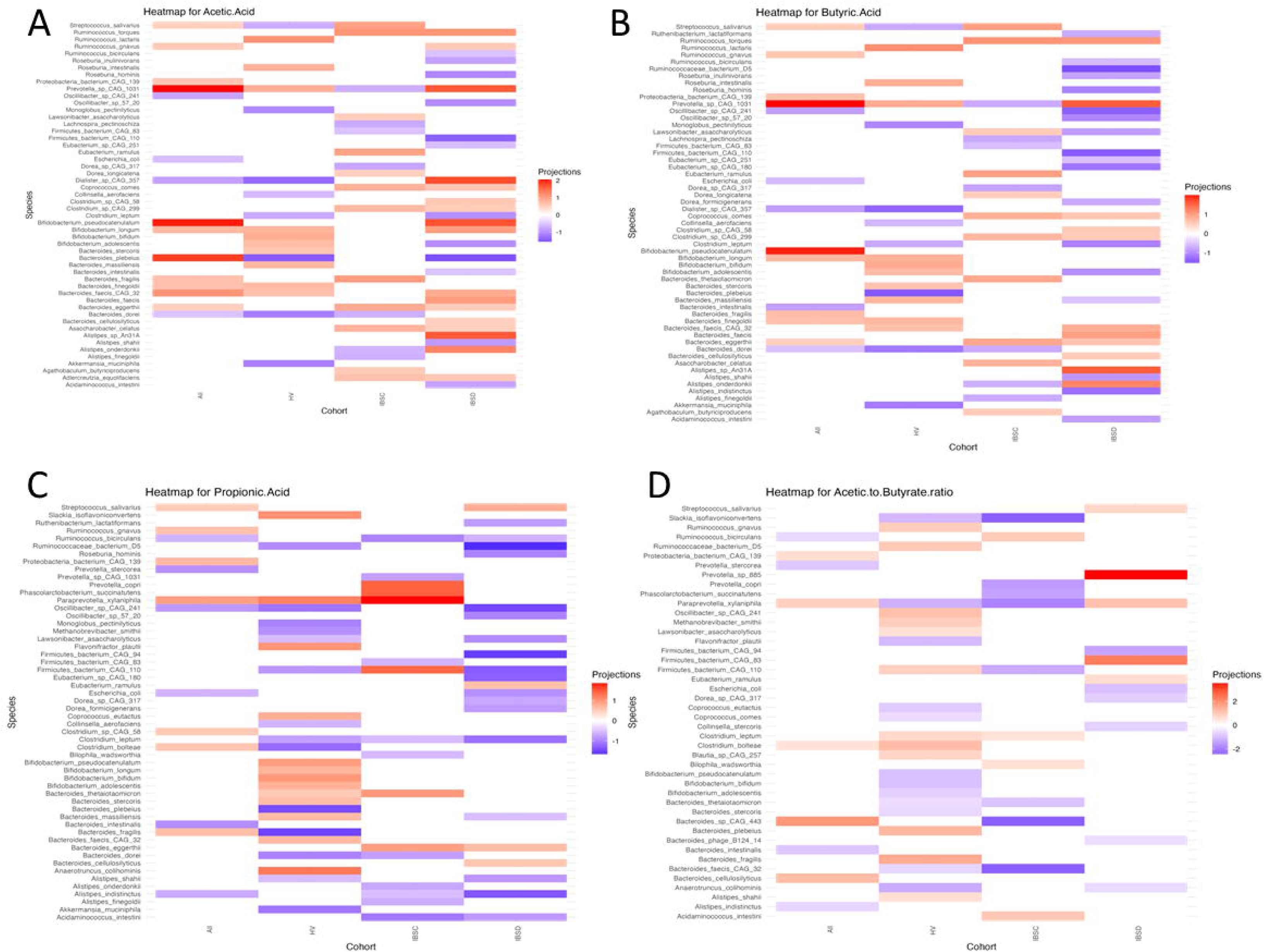
Correlations of bacterial taxa with short chain fatty acids (SCFA) based on partial canonical correspondence analysis. *Panels show associations between bacterial taxa and specific SCFA([a] acetate, [b] butyrate, [c] propionate, and [d] and acetate to butyrate ratio) within cohorts. The strength of the correlation is represented projections >0.5 with purple indicating negative correlations and red indicating positive correlations*.

In IBS-D, both positive and negative correlations with stool acetate were observed with multiple taxa. Largest negative projections were observed with several bacteria that have previously linked to SCFA production (*B. plebeius*, *Roseburia hominis*),^42, 43^ dietary polysaccharides and protein utilizers (*Firmicutes bacterium* CAG:110),^44^ starch or fructooligosaccharide degradation (*Bifidobacterium adolescentis*), fiber fermentation (e.g. *Clostridium leptum*^45^), β-fructan utilizers (e.g. *Roseburia inulinivorans*).^45^ Positive projections were observed with *Dialister* sp. CAG 357, *B. pseudocatenulatum*, *Alistipes* species, *Prevotella* sp. CAG:1031*, R. toques,* and *R. gnavus.* In IBS-C, largest negative projections for acetate were observed between abundances of *Dorea* sp. CAG:317, *Lachnospira pectinoschiza*, and *Prevotella sp.* CAG:1031 while the largest positive projections were observed with *Streptococcus salivarus*, *Bacteroides fragilis*, and *R. torques*. In controls, largest negative projections for acetate were observed with *B. plebeius, Dialister* sp. CAG:357, *Bacteroides dorei*, and *A. muciniphila*. The largest positive projection for acetate among controls was observed with *Ruminococcus lactaris*.

Analysis of microbe-butyrate associations in IBS-D demonstrated negative projection scores for butyrate with several bacteria including *Ruminococcaceae* bacterium D5, *Firmicutes bacterium* CAG:110, *C. leptum*, *R. hominis, R. inulinivorans*, and *L._asaccharolyticus*. The largest positive projection scores for butyrate were observed with *Alistipes* species, *Prevotella sp.* CAG:1031, and *R. torques*. In IBS-C and controls, patterns of microbe-butyrate associations largely mirrored the patterns of microbe-acetate associations observed within the respective clinical groups. In addition, a positive relationship between butyrate and *Bacteroides thetaiotaomicron*, a well-known human commensal^46, 47^ that exhibits the capacity to digest a broad array of polysaccharides and host glycans, was also observed in IBS-C.

For propionate, most microbes including *Ruminococcaceae* bacterium D5, *Firmicutes bacterium* CAG:110, *C. leptum*, *L. asaccharolyticus*, *Ruthenibacterium lactatiformans* and *Bacteroides massiliensis* exhibited negative associations in IBS-D with only a few exceptions including *S. salivarus*, for which the largest positive correlation was observed. In contrast, both positive and negative projections for stool propionate were observed in IBS-C; the largest positive scores were observed for *Paraprevotella xylaniphila*, *Phascolarctobacterium succinatutens*, and *P. copri*. Among controls, both positive and negative associations with propionate were observed for multiple bacterial taxa.

### Lower number of bacterial species associated with acetate to butyrate ratios in IBS

Since interconversion of SCFA from acetate to butyrate may represent a major route of butyrate formation within the gastrointestinal tract,^48^ we examined bacterial relationships with acetate to butyrate ratios to identify top taxa associated the metabolic processes that drive SCFA output. The largest positive associations were observed with *Prevotell*a sp. 885, *Firmicutes* bacterium CAG:83, *P. xylaniphila* in IBS-D, suggesting that these species were associated with shift toward decreased butyrate production. Unlike the patterns observed with individual SCFA, analysis acetate to butyrate ratios demonstrated that the number of ranked bacteria were substantially lower in IBS-D or IBS-C compared to controls (Figure 5), suggesting the possibility of reduced functional redundancy for butyrate formation in IBS.

### Reduced SCFA-producing microbes and bile acid malabsorption (BAM)

Stool microbial metagenomes were analyzed in 22 IBS-D patients with (n=8) and without (n=14) clinical BAM according to previously validated diagnostic cutoff values.^12, 13^ Comparisons of beta diversity based on Euclidian dissimilarity of ALR transformed abundance data showed significant dissimilarity (p = 0.039; R^2^ = 0.061) between patients with and without BAM (Supplemental Figure 3). Significant differences in six high abundant species were observed between groups, including *L. asaccharolyticus R. inulinivorans*, *L. pectinoschiza*, and *F. saccharivorans* which were also associated with stool SCFA, but not bile acids, in IBS-D. These four species with known SCFA-producing capacity were negatively correlated with BAM.

### Functional potential for carbohydrate degradation, SCFA metabolism, and mucin degradation

Examination of microbial gene families and pathways yielded 949,223 total gene families within the metagenome data set including 4,042 named KO terms. We applied a gut metabolic module (GMM) framework to analyze modules related to carbohydrate degradation, SCFA production or metabolism including bacterial cross-feeding pathways, mucin degradation, and one module related to serine degradation based on significant species-based predictors. Abundances of KO identifiers within modules for lactose degradation, (p=0.0026), serine degradation (p=0.019), and propionate production (p=0.026) were significantly enriched in IBS-C compared to controls. Relative abundance of a KO identifier within a galactose degradation module was significantly reduced in IBS-D (p=0.033) compared to controls. In the collective cohort, we applied the GMM framework to analyze the relationships between metagenomically-encoded functions and stool SCFA. We identified 23, 23, 15, and 16 KO identifiers from relevant GMMs that were significantly associated with total SCFA, acetate, butyrate, and propionate, respectively. Within a representative subset (n=6 controls, n=5 IBS-D, n=5 IBS-C) selected by stool form characteristics, functional metagenomic analyses demonstrated that microbial genes/pathways associated with SCFA production/metabolism and degradation of carbohydrates/mucin were differentially associated with clinical group (Figure 6).

**Figure 6:**
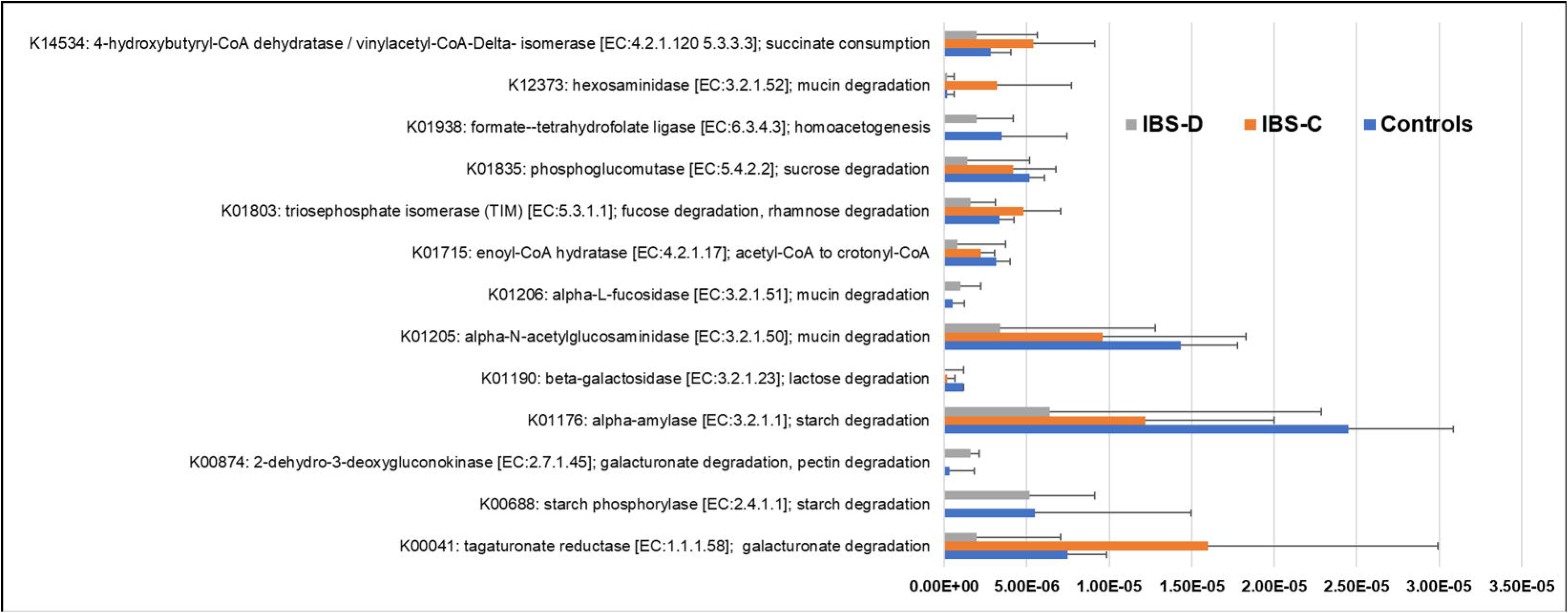
Variations in Microbial Substrate Utilization in Healthy Volunteers and Participants with IBS. *Figure shows significant differences in functional potential for carbohydrate degradation, short chain fatty acid production or metabolism, and mucin degradation across clinical groups in representative subset selected by stool form characteristics. Groups are denoted by color (HV = blue, IBS-C = red, IBS-D = gray)*.

## DISCUSSION

It has been proposed that changes in intestinal SCFAs in some patients with IBS are caused by shifts in microbial composition and function to drive and maintain symptoms. We hypothesized that specific features of the GI microbiome may correlate with SCFA output and that these relationships may distinguish IBS subtypes and endophenotypes. We further hypothesized that functional features of the GI microbiome defined by substrate preferences and SCFA metabolism may be linked to bowel dysfunction. Using a dual-omics approach, our findings demonstrate microbiota-SCFA relationships differ between IBS subtypes and endophenotypes, and between patients with IBS and healthy controls. We elected to focus on microbe-SCFA relationship due to the established biological relevance of SCFA in GI physiology and to use a focused strategy for identifying the microbes involved in shaping the microbial metabolome. Our findings suggest that key taxa may predict IBS mechanisms and that the functional repertoire of the intestinal microbiome influences bowel functions. By capturing quantifiable traits, we also gather evidence suggesting the microbiome has direct effects on excreted SCFA and were able to identify microbe-SCFA relationships that are not explained by transit, to potentially isolate major microbial features (or keystone taxa) that shape SCFA output.

While numerous studies have reported compositional changes^11^ in IBS such as decreased microbial diversity and altered abundances of individual taxa, consistent patterns are difficult to pinpoint and limit our ability to draw actional insights. Recent data suggest that putative microbial biomarkers are related to mechanistic and host-specific features in IBS rather than the overall clinical syndrome. Consistent with these prior studies, we observed differences in overall microbial community composition across groups. Comparisons of taxa abundances demonstrated significant changes were most common in IBS-D and predominantly characterized by higher abundances of several bacterial species including *Blautia* sp. CAG:257*, R. gnavus*, *Dorea* sp. CAG:317, and *Proteobacteria* bacterium CAG:139. *R. gnavus*, a Lachnospiraceae, has previously been identified to play a pathogenic role in IBS-D through serotonin biosynthesis,^41^ biofilm formation and increased stool bile acid excretion,^21^ production of proinflammatory polysaccharides,^49^ and mucin degradation.^50^ Interestingly, the relationship between *R. gnavus* and IBS lost its significant in adjusted analyses due to a positive association between *R. gnavus* and BMI which may suggest a role *R. gnavus* in explaining the connection between IBS-D or chronic diarrhea with metabolic syndrome and obesity-related disorders although further validation of this hypothesis will be required.^51^ Associations of IBS-D with other taxa including *Dorea* sp. CAG:317, *Blautia* sp. CAG:257, and *Proteobacteria* bacterium CAG:139 remained and a new association between *B. pseudocatenulatum* and IBS-D emerged. In our analyses of microbe-SCFA relationships, strongest contributions to SCFA in the overall cohort were observed with *Bacteroides plebeius, Prevotella sp.* CAG:1031, and *B. pseudocatenulatum* suggesting these taxa as major drivers of microbial metabolism which is consistent with their known functions. *B. plebeius* harbors a specialized polysaccharide utilization locus acquired through horizontal gene transfer from marine bacteria.^52, 53^ *Prevotella* spp. are highly prevalent within the human gut, and is largely regarded as a fiber degrader that is associated with agrarian or plant-based diets, although strain-level variations may impact their contributions to health and disease.^54^ *B. pseudocatenulatum* is recognized as an adult-type *Bifidobacterium* that is metabolically adapted toward dietary carbohydrates including long-chain xylans.^55^ Among the three clinical groups, we found microbe-SCFA associations were most pronounced in IBS-D and that fewer microbes were linked to acetate and butyrate ratios. These observations may indicate that microbial contributions to SCFA profiles are particularly important in IBS-D and may also imply a greater degree of microbial specialization and reduced functional redundancy. Previously, Jacobs et al.^56^ reported significant increases in transcript abundances for fructooligosaccharide and polyol utilization and upregulation of transcripts for fructose and glucan metabolism as well as the succinate pathway for carbohydrate fermentation in IBS. Comparisons between IBS-D and IBS-C demonstrated differences in microbial metabolism with upregulation of multiple metabolic pathways including transcripts for fructose, mannose, and polyol metabolism in IBS-D. Together, our findings suggest that differences microbial metabolism between IBS subtypes may underlie variations in clinical symptoms or that changes in microbial metabolism could play a particularly prominent role in IBS-D. These findings could explain the higher prevalence of mixed- and diarrhea-predominant symptoms in post-infection IBS^57, 58^ as well as the limited evidence for efficacy of antimicrobial treatments such as rifaximin in IBS-C^59^ relative to IBS-D.^60, 61^

*Dorea* sp. CAG:317 and *B. pseudocatenulatum* were identified as top ranked bacteria associated with acetate to butyrate ratio and acetate concentrations. Mucin degradation has been described among several *Dorea* species and the genus *Dorea* belongs to the Lachnospiraceae family, a major producer of SCFA,^62^ Notably, Wang et al.^63^ recently demonstrated that while xylan supplementation following fiber deprivation alleviated gut dysbiosis through selective promotion of *B. pseudocatenulatum*, these changes were found to be associated with lower community diversity. *Dorea* sp. CAG:317 and *B. pseudocatenulatum* may represent keystone taxa linked to altered microbial metabolism or persistent impairment in community recovery among patients with IBS-D. Only two bacterial species, *A. muciniphila* and *P. copri* were significantly increased in IBS-C. Although relationships between microbes and SCFA were less clear in IBS-C. *P. copri* was further identified as top ranked bacteria that was positively associated with propionate and negatively associated with acetate to butyrate ratio in IBS-C. Previous studies have reported increased *A. muciniphila* in IBS-C to be protective against experimentally-induced colitis in mice^64^ and a positive association between mucosal *P. copri* abundance and abdominal pain in patients with IBS without prior history of infection.^65^ Others have demonstrated that IBS patients with high levels of propionate present with worse gastrointestinal symptoms, quality of life, and negative emotions.^15^ *P. copri* is a prominent gut commensal that contributes to propionate production via the succinate pathway^66^ and has been linked to both beneficial and harmful effects on human health. Rolhion et al. found *P. copri* to be associated with mucus erosion and increased propionate. In another study by Jiang et. al, *P. copri* colonization was associated with proinflammatory effects following a a high fiber diet in patients with rheumatoid arthritis through succinate production.^67^ Exacerbation of rheumatoid with the high fiber diet in the presence of *P. copri* colonization led to microbial dysbiosis characterized by increased abundance of *Akkermansia*, which was also increased in our own IBS-C cohort.

In addition to identify major microbial features associated with individual SCFA, we made several important observations related to the pattern or direction of observed microbe-SCFA relationships. Interestingly, negative correlations were observed in the IBS-D subgroup between several SCFA-producing species including *R. hominis, B. adolescentis*, *R. inulinivorans* and individual SCFA. *B. adolescentis* utilizes fructo-oligosaccharides and starch and leads to the production of lactate and acetate which is cross-fed to butyrate-producing anaerobes,^68^ but have also been identified as a potential GABA producer and correlated with anxiety and/or depression,^69^ which are common comorbid conditions in patients with IBS. *R. inulinivorans* is a known fiber fermenter^45^ that also possesses the ability to forage mucin through a metabolic interplay with other members of the intestinal microbiota^70^ and is also known for net acetate uptake,^66^ which may explain its correlation with reduced SCFA. In general, analysis of bacterial associations with individual SCFAs demonstrated the largest number of negative associations in IBS-D. Many of these negative correlations involved bacterial species that have been implicated in SCFA production and polysaccharide utilization. Findings may imply disrupted cross-feeding mechanisms, modifications in substrate utilization, competitive inhibition, or other context-specific changes leading to altered microbial metabolism in IBS. Upon analyzing relationships of bacterial species with acetate to butyrate ratios, which we studied as a marker for overall SCFA production and intestinal SCFA availability,^71^ we identified fewer top ranked bacteria in IBS compared to healthy controls to suggest a reduction in overall functional redundancy within the IBS microbiome. Future work should investigate the effects of bacterial cross-feeding, microbial conversions, and interactions of the colonic microbiome with diet- and host-derived substrates that may destabilize the microbiota-mucosal interface to expose vulnerabilities and promote pathophysiological mechanisms that drive IBS.

A few bacterial species were associated with SCFA in both IBS subtypes including *R. torques*, *C. comes*, *Clostridium* sp. CAG:299, *B. eggerthii*, and *Adlercreutzia equolifaciens* which may suggest these bacterial species represent shared microbial features across IBS subtypes. *R. torques* is a mucin-degrading bacteria that has previously been associated with IBS^72^ as well as depression.^73^ Similarly, recent studies have demonstrated associations between *C. comes*^74^ and *B. eggerthii*^75^ depression and PTSD, which have both been described as comorbid conditions in IBS. *R. A. equolifaciens* has been described as an anti-inflammatory commensal in non-alcoholic fatty liver disease.^76^ These data could point towards shared microbial features as novel biomarker of common pathogenic mechanisms in IBS involving the microbiota-gut-brain axis, possibly through the effects of microbially-derived SCFA.

In our comparisons across IBS-D patients by with and without BAM, we observed BAM to be associated with reduced abundances of several SCFA-producing bacterial species. Similar observations have been made in studies linking biofilm formation to bile acid accumulation and reduced SCFA-producing bacteria.^21^ Results suggest that BAM exerts indirect effects on the intestinal metabolome or the overall metabolic potential of the colonic microbiota characterized by a decline in SCFA-producing capacity. We also assessed of functional variations in the GI microbiome to reveal differential abundances of KO identifiers belonging to metabolic processes related to degradation of carbohydrates and serine as well as SCFA production. In a subset of participants selected by stool form characteristics, we identified changes in substrate degradation potential and fermentative capacity between IBS and controls and between IBS subtypes that were characterized by increased capacity for starch degradation in controls, enhancement of distinct mucin-degrading functions in IBS-D vs. IBS-C, and reduced pectin degrading potential in IBS-C. Results suggest that abnormal stool form in IBS is influenced by microbiota-encoded substrate preferences.

Collectively, our findings indicate there are distinct changes in microbiome composition may underlie clinical heterogeneity in IBS. Using a dual-omics approach, we demonstrate that the gut microbiota in IBS-D is characterized by increased metabolic activity, reduced functional redundancy, and persistently reduced diversity. Several taxa including *R. gnavus*, *Dorea* sp. CAG:317 and *B. pseudocatenulatum* are identified as the leading candidates for further study as potential microbial targets in IBS-D. A few unique microbial features including *P. copri*-associated propionate production and *Akkermansia* expansion may play a crucial role in IBS-C. Meanwhile, microbial features that are common between subtypes may offer insights into shared pathophysiological mechanisms involving the microbiota-gut-brain axis in IBS. Identifying the major microbial features (taxa or functions) that drive metabolic output may be crucial in defining rational approaches to modulate the microbiome in IBS.

Despite study strengths, we recognize the limitations of this work including cross-sectional sampling, which overlooks the fluctuating nature of the microbiome. To address this, we assessed relationships between microbial abundances and quantifiable IBS endophenotypes that were defined at the time of specimen collection while accounting for baseline covariates. All participants were instructed to consume a 4-day high fat diet while otherwise maintaining their usual diet. Previously, we examined the effect of both habitual diet and real-time intake on excreted stool SCFA to find that that modest variations in macronutrient intake including polysaccharides did not exert substantial effects on excreted SCFA.^25, 77^ We enrolled patients with opposing IBS phenotypes from both the university clinics and the surrounding communities to increase our ability to detect differences between clinical cohorts and limit referral bias. We also acknowledge that causal microbial mechanisms cannot be determined through the current work. However, we applied a dual-omics approach to investigate biologically relevant changes in intestinal microbiome composition and function. Finally, we cannot discount the possibility we may have missed significant taxa due to the moderate sample size and a risk for Type I errors as we did not adjust for false discovery rate in our analysis of taxonomic differences. However, our analytic approach was based on endpoints that were determined *a priori* and we applied a conservative strategy by focusing on taxa that were associated with SCFA while accounting for transit. We further limited analyses to high abundant taxa and to species that exhibited >3-fold change in abundance while focusing on the biological plausibility of our results. Findings from this study should be validated in larger, longitudinal cohorts, but provide a framework that may be used to define key microbial interactions and targets in IBS.

In conclusion, main findings from this study highlight microbiota-SCFA patterns vary across clinical IBS phenotypes and endophenotypes. Our observations suggest that a paired microbe-metabolite approach may represent a viable strategy for identifying actionable microbial features in heterogenous patient populations. Prominent shifts in microbial composition are observed in IBS-D. These changes appear to affect metabolic capacity, substrate preferences, and metabolite profiles. In addition, altered microbial substrate uptake may impact stool form or bowel function in IBS. These findings generate new hypotheses regarding candidate microbes that could be pursued as actionable microbial biomarkers or metabolically influential features in the effort to develop rational microbiota-based therapies in IBS.

## Data Availability

Microbiome sequencing reads are deposited into NCBI with accession number SUB13882354. The data that support the findings of this study are available upon reasonable request from the corresponding author.

## Acknowledgements

The authors would like to thank the Mayo Clinic Department of Laboratory Medicine and Pathology and Purdue University Metabolite Profiling Facility for their collaboration and assistance with quantification of stool bile acids and short chain fatty acids.

## Abbreviations

ALR: additive log-ratio
BAM: bile acid malabsorption
CTT: colonic transit time
gDNA: genomic DNA
GI: gastrointestinal
GLM: generalized linear model
GMM: gut metabolic module
IBS: irritable bowel syndrome
IBS-D: IBS with diarrhea
IBS-C: IBS with constipation
KEGG: Kyoto Encyclopedia of Genes and Genomics
KO: KEGG Orthogroup
LC-MS: liquid chromatography-mass spectrometry
SCFA: short chain fatty acids

## Author Contributions

Developing study concept: AS. Planning study design: AS, HX, XG. Participant recruitment: AS, MRW, RS, TJS, NR, MB JW, CL, AG, JK, RA. Data collection and study procedures: AS, MRW, CL, AG, JK, ET. Data Management: AS, YX, MRW, CL, JK, HX, XG. Data Analysis and Interpretation: YX, XG. Drafting the manuscript: AS, XG. Critically revising the manuscript: MRW, RS, TJS, NR, MB, JW, ET, HX.

## References

1. Sperber AD, Bangdiwala SI, Drossman DA, et al. Worldwide Prevalence and Burden of Functional Gastrointestinal Disorders, Results of Rome Foundation Global Study. Gastroenterology 2021;160:99–114 e3.

2. Ford AC, Sperber AD, Corsetti M, Camilleri M. Irritable bowel syndrome. Lancet 2020;396:1675–1688.

3. Tap J, Derrien M, Tornblom H, et al. Identification of an Intestinal Microbiota Signature Associated With Severity of Irritable Bowel Syndrome. Gastroenterology 2017;152:111–123 e8.

4. Anitha M, Vijay-Kumar M, Sitaraman SV, et al. Gut microbial products regulate murine gastrointestinal motility via Toll-like receptor 4 signaling. Gastroenterology 2012;143:1006–16 e4.

5. Heitkemper MM, Cain KC, Shulman RJ, et al. Stool and urine trefoil factor 3 levels: associations with symptoms, intestinal permeability, and microbial diversity in irritable bowel syndrome. Benef Microbes 2018;9:345–355.

6. Sundin J, Rangel I, Repsilber D, Brummer RJ. Cytokine Response after Stimulation with Key Commensal Bacteria Differ in Post-Infectious Irritable Bowel Syndrome (PI-IBS) Patients Compared to Healthy Controls. PLoS One 2015;10:e0134836.

7. McCracken VJ, Lorenz RG. The gastrointestinal ecosystem: a precarious alliance among epithelium, immunity and microbiota. Cell Microbiol 2001;3:1–11.

8. Liu Y, Zhang L, Wang X, et al. Similar Fecal Microbiota Signatures in Patients With Diarrhea-Predominant Irritable Bowel Syndrome and Patients With Depression. Clin Gastroenterol Hepatol 2016;14:1602–1611 e5.

9. Crouzet L, Gaultier E, Del’Homme C, et al. The hypersensitivity to colonic distension of IBS patients can be transferred to rats through their fecal microbiota. Neurogastroenterol Motil 2013;25:e272–82.

10. Mars RAT, Frith M, Kashyap PC. Functional Gastrointestinal Disorders and the Microbiome-What Is the Best Strategy for Moving Microbiome-based Therapies for Functional Gastrointestinal Disorders into the Clinic? Gastroenterology 2021;160:538–555.

11. Pittayanon R, Lau JT, Yuan Y, et al. Gut Microbiota in Patients With Irritable Bowel Syndrome-A Systematic Review. Gastroenterology 2019;157:97–108.

12. Shin A, Camilleri M, Vijayvargiya P, et al. Bowel functions, fecal unconjugated primary and secondary bile acids, and colonic transit in patients with irritable bowel syndrome. Clin Gastroenterol Hepatol 2013;11:1270–1275 e1.

13. Vijayvargiya P, Camilleri M, Burton D, et al. Bile and fat excretion are biomarkers of clinically significant diarrhoea and constipation in irritable bowel syndrome. Aliment Pharmacol Ther 2019;49:744–758.

14. Mars RAT, Yang Y, Ward T, et al. Longitudinal Multi-omics Reveals Subset-Specific Mechanisms Underlying Irritable Bowel Syndrome. Cell 2020;182:1460–1473 e17.

15. Tana C, Umesaki Y, Imaoka A, et al. Altered profiles of intestinal microbiota and organic acids may be the origin of symptoms in irritable bowel syndrome. Neurogastroenterol Motil 2010;22:512–9, e114-5.

16. Vijayvargiya P, Camilleri M, Chedid V, et al. Analysis of Fecal Primary Bile Acids Detects Increased Stool Weight and Colonic Transit in Patients With Chronic Functional Diarrhea. Clin Gastroenterol Hepatol 2019;17:922–929 e2.

17. BouSaba J, Sannaa W, McKinzie S, et al. Impact of Bile Acid Diarrhea in Patients With Diarrhea-Predominant Irritable Bowel Syndrome on Symptoms and Quality of Life. Clin Gastroenterol Hepatol 2022;20:2083–2090 e1.

18. BouSaba J, Zheng T, Dilmaghani S, et al. Effect of rapid colonic transit on stool microbiome and short-chain fatty acids in diarrhoea-predominant irritable bowel syndrome. Gut 2023.

19. Camilleri M, Carlson P, BouSaba J, et al. Comparison of biochemical, microbial and mucosal mRNA expression in bile acid diarrhoea and irritable bowel syndrome with diarrhoea. Gut 2023;72:54–65.

20. Zhao L, Yang W, Chen Y, et al. A Clostridia-rich microbiota enhances bile acid excretion in diarrhea-predominant irritable bowel syndrome. J Clin Invest 2020;130:438–450.

21. Baumgartner M, Lang M, Holley H, et al. Mucosal Biofilms Are an Endoscopic Feature of Irritable Bowel Syndrome and Ulcerative Colitis. Gastroenterology 2021;161:1245–1256 e20.

22. Silva YP, Bernardi A, Frozza RL. The Role of Short-Chain Fatty Acids From Gut Microbiota in Gut-Brain Communication. Front Endocrinol (Lausanne) 2020;11:25.

23. Ringel-Kulka T, Choi CH, Temas D, et al. Altered Colonic Bacterial Fermentation as a Potential Pathophysiological Factor in Irritable Bowel Syndrome. Am J Gastroenterol 2015;110:1339–46.

24. Gargari G, Taverniti V, Gardana C, et al. Fecal Clostridiales distribution and short-chain fatty acids reflect bowel habits in irritable bowel syndrome. Environ Microbiol 2018;20:3201–3213.

25. Waseem MR, Shin A, Siwiec R, et al. Associations of Fecal Short Chain Fatty Acids With Colonic Transit, Fecal Bile Acid, and Food Intake in Irritable Bowel Syndrome. Clin Transl Gastroenterol 2023;14:e00541.

26. Deehan EC, Yang C, Perez-Munoz ME, et al. Precision Microbiome Modulation with Discrete Dietary Fiber Structures Directs Short-Chain Fatty Acid Production. Cell Host Microbe 2020;27:389–404 e6.

27. Mearin F, Lacy BE, Chang L, et al. Bowel Disorders. Gastroenterology 2016.

28. Mulligan AA, Luben RN, Bhaniani A, et al. A new tool for converting food frequency questionnaire data into nutrient and food group values: FETA research methods and availability. BMJ Open 2014;4:e004503.

29. Heaton KW, Radvan J, Cripps H, et al. Defecation frequency and timing, and stool form in the general population: a prospective study. Gut 1992;33:818–24.

30. Sadik R, Abrahamsson H, Ung KA, Stotzer PO. Accelerated regional bowel transit and overweight shown in idiopathic bile acid malabsorption. Am J Gastroenterol 2004;99:711–8.

31. Park J, Kim M, Kang SG, et al. Short-chain fatty acids induce both effector and regulatory T cells by suppression of histone deacetylases and regulation of the mTOR-S6K pathway. Mucosal Immunol 2015;8:80–93.

32. Tagliacozzi D, Mozzi AF, Casetta B, et al. Quantitative analysis of bile acids in human plasma by liquid chromatography-electrospray tandem mass spectrometry: a simple and rapid one-step method. Clin Chem Lab Med 2003;41:1633–41.

33. Vijayvargiya P, Camilleri M, Shin A, Saenger A. Methods for diagnosis of bile acid malabsorption in clinical practice. Clin Gastroenterol Hepatol 2013;11:1232–9.

34. Louis P, Hold GL, Flint HJ. The gut microbiota, bacterial metabolites and colorectal cancer. Nat Rev Microbiol 2014;12:661–72.

35. McMurdie PJ, Holmes S. Phyloseq: a bioconductor package for handling and analysis of high-throughput phylogenetic sequence data. Pac Symp Biocomput 2012:235–46.

36. Oksanen J, Simpson GL, Blanchet F, et al. vegan: Community Ecology Package. Ordination methods, diversity analysis and other functions for community and vegetation ecologists. 2.6-4 ed, 2022.

37. Vieira-Silva S, Falony G, Darzi Y, et al. Species-function relationships shape ecological properties of the human gut microbiome. Nat Microbiol 2016;1:16088.

38. Vujkovic-Cvijin I, Sklar J, Jiang L, et al. Host variables confound gut microbiota studies of human disease. Nature 2020;587:448–454.

39. Saad RJ, Rao SS, Koch KL, et al. Do stool form and frequency correlate with whole-gut and colonic transit? Results from a multicenter study in constipated individuals and healthy controls. Am J Gastroenterol 2010;105:403–11.

40. Jaruvongvanich V, Patcharatrakul T, Gonlachanvit S. Prediction of Delayed Colonic Transit Using Bristol Stool Form and Stool Frequency in Eastern Constipated Patients: A Difference From the West. J Neurogastroenterol Motil 2017;23:561–568.

41. Zhai L, Huang C, Ning Z, et al. Ruminococcus gnavus plays a pathogenic role in diarrhea-predominant irritable bowel syndrome by increasing serotonin biosynthesis. Cell Host Microbe 2023;31:33–44 e5.

42. Yun EJ, Yu S, Park NJ, et al. Metabolic and enzymatic elucidation of cooperative degradation of red seaweed agarose by two human gut bacteria. Sci Rep 2021;11:13955.

43. Machiels K, Joossens M, Sabino J, et al. A decrease of the butyrate-producing species Roseburia hominis and Faecalibacterium prausnitzii defines dysbiosis in patients with ulcerative colitis. Gut 2014;63:1275–83.

44. Quan J, Wu Z, Ye Y, et al. Metagenomic Characterization of Intestinal Regions in Pigs With Contrasting Feed Efficiency. Front Microbiol 2020;11:32.

45. Armstrong HK, Bording-Jorgensen M, Santer DM, et al. Unfermented beta-fructan Fibers Fuel Inflammation in Select Inflammatory Bowel Disease Patients. Gastroenterology 2023;164:228–240.

46. Martens EC, Chiang HC, Gordon JI. Mucosal glycan foraging enhances fitness and transmission of a saccharolytic human gut bacterial symbiont. Cell Host Microbe 2008;4:447–57.

47. Porter NT, Luis AS, Martens EC. Bacteroides thetaiotaomicron. Trends Microbiol 2018;26:966–967.

48. Shetty SA, Boeren S, Bui TPN, et al. Unravelling lactate-acetate and sugar conversion into butyrate by intestinal Anaerobutyricum and Anaerostipes species by comparative proteogenomics. Environ Microbiol 2020;22:4863–4875.

49. Henke MT, Kenny DJ, Cassilly CD, et al. Ruminococcus gnavus, a member of the human gut microbiome associated with Crohn’s disease, produces an inflammatory polysaccharide. Proc Natl Acad Sci U S A 2019;116:12672–12677.

50. Bell A, Brunt J, Crost E, et al. Elucidation of a sialic acid metabolism pathway in mucus-foraging Ruminococcus gnavus unravels mechanisms of bacterial adaptation to the gut. Nat Microbiol 2019;4:2393–2404.

51. Shin A, Xu H, Imperiale TF. Associations of chronic diarrhoea with non-alcoholic fatty liver disease and obesity-related disorders among US adults. BMJ Open Gastroenterol 2019;6:e000322.

52. Hehemann JH, Correc G, Barbeyron T, et al. Transfer of carbohydrate-active enzymes from marine bacteria to Japanese gut microbiota. Nature 2010;464:908–12.

53. Hehemann JH, Kelly AG, Pudlo NA, et al. Bacteria of the human gut microbiome catabolize red seaweed glycans with carbohydrate-active enzyme updates from extrinsic microbes. Proc Natl Acad Sci U S A 2012;109:19786–91.

54. De Filippis F, Pasolli E, Tett A, et al. Distinct Genetic and Functional Traits of Human Intestinal Prevotella copri Strains Are Associated with Different Habitual Diets. Cell Host Microbe 2019;25:444–453 e3.

55. Derrien M, Turroni F, Ventura M, van Sinderen D. Insights into endogenous Bifidobacterium species in the human gut microbiota during adulthood. Trends Microbiol 2022;30:940–947.

56. Jacobs JP, Lagishetty V, Hauer MC, et al. Multi-omics profiles of the intestinal microbiome in irritable bowel syndrome and its bowel habit subtypes. Microbiome 2023;11:5.

57. Klem F, Wadhwa A, Prokop LJ, et al. Prevalence, Risk Factors, and Outcomes of Irritable Bowel Syndrome After Infectious Enteritis: A Systematic Review and Meta-analysis. Gastroenterology 2017;152:1042–1054 e1.

58. Breen-Lyles M, Decuir M, Byale A, et al. Impact of Rome IV criteria on the prevalence of post-infection irritable bowel syndrome. Neurogastroenterol Motil 2023;35:e14532.

59. Pimentel M, Chang C, Chua KS, et al. Antibiotic treatment of constipation-predominant irritable bowel syndrome. Dig Dis Sci 2014;59:1278–85.

60. Pimentel M, Lembo A, Chey WD, et al. Rifaximin therapy for patients with irritable bowel syndrome without constipation. N Engl J Med 2011;364:22–32.

61. Lembo A, Sultan S, Chang L, et al. AGA Clinical Practice Guideline on the Pharmacological Management of Irritable Bowel Syndrome With Diarrhea. Gastroenterology 2022;163:137–151.

62. Vacca M, Celano G, Calabrese FM, et al. The Controversial Role of Human Gut Lachnospiraceae. Microorganisms 2020;8.

63. Wang Z, Bai Y, Pi Y, et al. Xylan alleviates dietary fiber deprivation-induced dysbiosis by selectively promoting Bifidobacterium pseudocatenulatum in pigs. Microbiome 2021;9:227.

64. Gobert AP, Sagrestani G, Delmas E, et al. The human intestinal microbiota of constipated-predominant irritable bowel syndrome patients exhibits anti-inflammatory properties. Sci Rep 2016;6:39399.

65. Choo C, Mahurkar-Joshi S, Dong TS, et al. Colonic mucosal microbiota is associated with bowel habit subtype and abdominal pain in patients with irritable bowel syndrome. Am J Physiol Gastrointest Liver Physiol 2022;323:G134–G143.

66. Louis P, Flint HJ. Formation of propionate and butyrate by the human colonic microbiota. Environ Microbiol 2017;19:29–41.

67. Jiang L, Shang M, Yu S, et al. A high-fiber diet synergizes with Prevotella copri and exacerbates rheumatoid arthritis. Cell Mol Immunol 2022;19:1414–1424.

68. Belenguer A, Duncan SH, Calder AG, et al. Two routes of metabolic cross-feeding between Bifidobacterium adolescentis and butyrate-producing anaerobes from the human gut. Appl Environ Microbiol 2006;72:3593–9.

69. Duranti S, Ruiz L, Lugli GA, et al. Bifidobacterium adolescentis as a key member of the human gut microbiota in the production of GABA. Sci Rep 2020;10:14112.

70. Pichler MJ, Yamada C, Shuoker B, et al. Butyrate producing colonic Clostridiales metabolise human milk oligosaccharides and cross feed on mucin via conserved pathways. Nat Commun 2020;11:3285.

71. Wijdeveld M, Schrantee A, Hagemeijer A, et al. Intestinal acetate and butyrate availability is associated with glucose metabolism in healthy individuals. iScience 2023;26:108478.

72. Carco C, Young W, Gearry RB, et al. Increasing Evidence That Irritable Bowel Syndrome and Functional Gastrointestinal Disorders Have a Microbial Pathogenesis. Front Cell Infect Microbiol 2020;10:468.

73. Mulder D, Jakobi B, Shi Y, et al. Gut microbiota composition links to variation in functional domains across psychiatric disorders. Brain Behav Immun 2024;120:275–287.

74. Yu L, Chen X, Bai X, et al. Microbiota Alters and Its Correlation with Molecular Regulation Underlying Depression in PCOS Patients. Mol Neurobiol 2023.

75. Xiao L, Liu S, Wu Y, et al. The interactions between host genome and gut microbiome increase the risk of psychiatric disorders: Mendelian randomization and biological annotation. Brain Behav Immun 2023;113:389–400.

76. Onate FP, Chamignon C, Burz SD, et al. Adlercreutzia equolifaciens Is an Anti-Inflammatory Commensal Bacterium with Decreased Abundance in Gut Microbiota of Patients with Metabolic Liver Disease. Int J Mol Sci 2023;24.

77. Calderon G, Patel C, Camilleri M, et al. Associations of Habitual Dietary Intake With Fecal Short-Chain Fatty Acids and Bowel Functions in Irritable Bowel Syndrome. J Clin Gastroenterol 2022;56:234–242.

