## Supplemental Methods for "Microbiota-Short Chain Fatty Acid Relationships Underlie Clinical Heterogeneity and Identify Key Microbial Targets in Irritable Bowel Syndrome (IBS)"

**Participant Eligibility Criteria**

Individuals with a history of microscopic colitis, inflammatory bowel disease, celiac disease, visceral cancer, immunodeficiency, known liver disease or elevated AST/ALT >2.0x the upper limit of normal, radiation therapy of abdomen, uncontrolled hypothyroidism, pre- or probiotic use within the past two weeks, regular tobacco-use, antibiotic-use within the past three months, pregnant, or abdominal surgeries (caesarean-section, tubal ligation, vaginal hysterectomy, appendectomy, or cholecystectomy >6 months prior to study initiation) were excluded. Participants were not permitted to take over the counter or prescription medications that could affect GI transit or study interpretation (e.g., opioids, narcotics, anticholinergics, norepinephrine reuptake inhibitors, nonsteroidal anti-inflammatory agents, COX-2 inhibitors, bile acid sequestrants) within two days and six months of study initiation for IBS and control participants, respectively. Participants were on stable diet regimens and not following modified diets for IBS symptoms (e.g., gluten-free or diet low in fermentable oligo-, di-, monosaccharides, and polyols) at the time of enrollment.
