## Supplemental Figure 1 for "Microbiota-Short Chain Fatty Acid Relationships Underlie Clinical Heterogeneity and Identify Key Microbial Targets in Irritable Bowel Syndrome (IBS)"

**Supplemental Figure 1:** β-Diversity Ordination of Samples from Health Control and Irritable Bowel Syndrome (IBS) Participants Including IBS with Constipation (IBS-C) and IBS with Diarrhea (IBS-D)

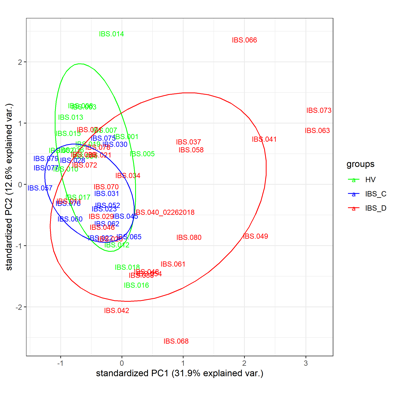
