## Supplemental Figure 2 for "Microbiota-Short Chain Fatty Acid Relationships Underlie Clinical Heterogeneity and Identify Key Microbial Targets in Irritable Bowel Syndrome (IBS)"

**Supplemental Figure 2:** Partial Canonical Correspondence Analysis on Microbiome and Short Chain Fattty Acid Data, Conditioned on Transit Time, in Individual Clinical Groups

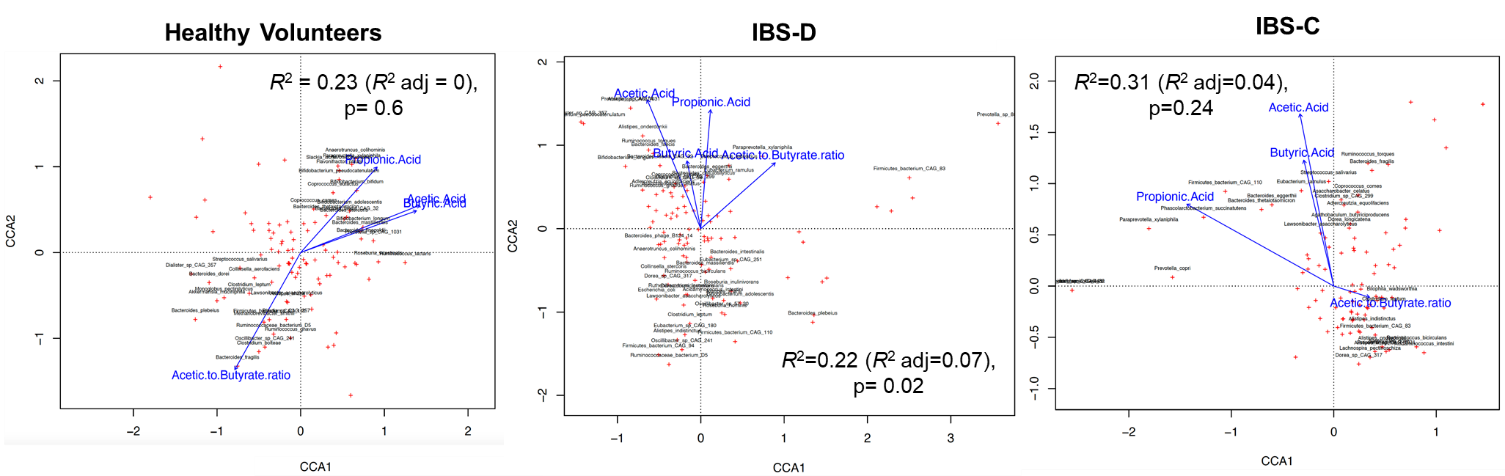
