## Supplemental Figure 3 for "Microbiota-Short Chain Fatty Acid Relationships Underlie Clinical Heterogeneity and Identify Key Microbial Targets in Irritable Bowel Syndrome (IBS)"

**Supplemental Figure 3:** Non-metric multidimensional scaling (NMDS) representation of beta diversity in patients with (yes) and without (no) bile acid malabsorption (BAM) using taxonomic classification from MetaPhlAn.

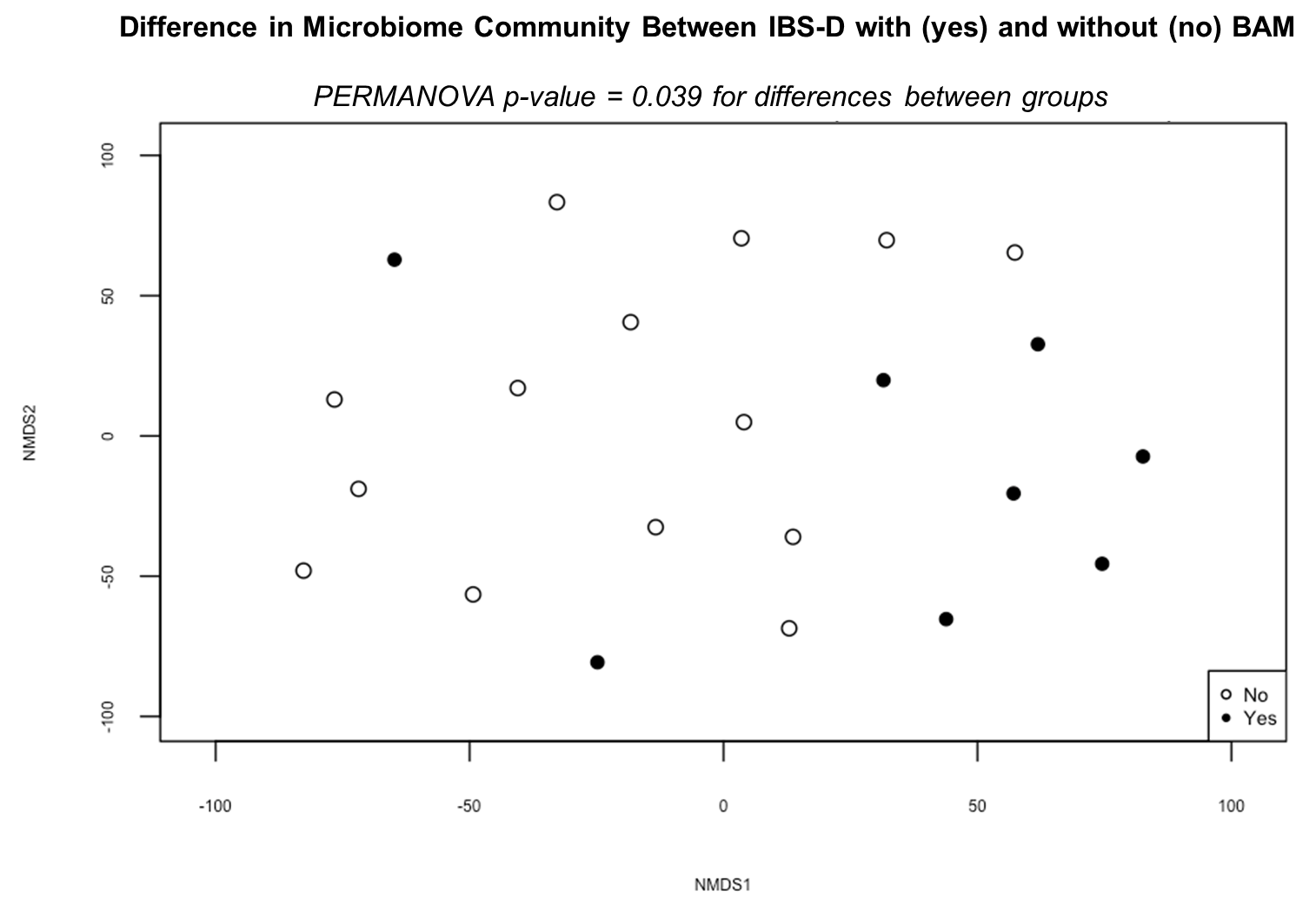
